# Mental and Emotional Health of Youth after 24 months of Gender-Affirming Medical Care Initiated with Pubertal Suppression

**DOI:** 10.1101/2025.05.14.25327614

**Authors:** Johanna Olson-Kennedy, Ramon Durazo-Arvizu, Liyuan Wang, Carolyn F. Wong, Diane Chen, Diane Ehrensaft, Marco A. Hidalgo, Yee-Ming Chan, Robert Garofalo, Asa E. Radix, Stephen M. Rosenthal

**Affiliations:** Division of Adolescent and Young Adult Medicine, Children’s Hospital, Los Angeles, California; Department of Pediatrics, University of Southern California, Los Angeles, California; The Saban Research Institute, Children’s Hospital Los Angeles; Division of Research on Children, Keck School of Medicine of USC; Potocsnak Family Division of Adolescent and Young Adult Medicine, Ann & Robert H. Lurie Children’s Hospital of Chicago, Chicago, Illinois; Pritzker Department of Psychiatry and Behavioral Health, Ann & Robert H. Lurie Children’s Hospital of Chicago, Chicago, Illinois; Department of Pediatrics, Northwestern University Feinberg School of Medicine, Chicago, Illinois; Department of Psychiatry and Behavioral Sciences, Northwestern University Feinberg School of Medicine, Chicago, Illinois; Department of Medical Social Sciences, Northwestern University Feinberg School of Medicine, Chicago, Illinois; Child and Adolescent Gender Center, Department of Pediatrics, University of California San Francisco, San Francisco, California; Department of Pediatrics, UCSF Benioff Children’s Hospital, San Francisco, California; Division of General Internal Medicine and Health Services Research, Department of Medicine, David Geffen School of Medicine at UCLA, Los Angeles, California (previously a,b); Division of Endocrinology, Boston Children’s Hospital, Boston, Massachusetts; Department of Pediatrics, Harvard Medical School, Boston, Massachusetts; Department of Medicine, Callen-Lorde Community Health Center, New York, New York; Department of Epidemiology, Columbia University Mailman School of Public Health, New York, New York

**Author notes:** **Address correspondence to**: Johanna Olson-Kennedy, MD, Children’s Hospital Los Angeles, 4650 Sunset Blvd. MS 2, Los Angeles, CA 90027. **Clinical Trial Registration (if any):** none.

## Abstract

**Background and Objectives:** Medical interventions for youth with gender dysphoria can include the use of gonadotropin releasing hormone analogs (GnRHas) for suppression of endogenous puberty. This analysis aimed to understand the impact of medical intervention initiated with GnRHas on psychological well-being among youth with gender dysphoria over 24 months.

**Methods:** Participants were enrolled as part of the Trans Youth Care United States Study. Eligibility criteria for youth included a diagnosis of Gender Dysphoria and pubertal initiation. Youth with precocious puberty or pre-existing osteoporosis were ineligible. Youth reported on depressive symptoms, emotional health and suicidality at baseline, 6, 12, 18 and 24 months after initiation of GnRHas. Parent/caretaker completed the Child Behavior Checklist at baseline, 12 and 24 months after initiation of GnRHas. Latent Growth-Curve Models analyzed trajectories of change over the 24-month period.

**Results:** Ninety-four youth aged 8-16 years (mean=11.2 y) were predominately Non-Hispanic White (56%), early pubertal (86%) and assigned male at birth (52%). Depression symptoms, emotional health and CBCL constructs did not change significantly over 24 months. At no time points were the means of depression, emotional health or CBCL constructs in a clinically concerning range.

**Conclusion:** Participants initiating medical interventions for gender dysphoria with GnRHas have self- and parent-reported psychological and emotional health comparable with the population of adolescents at large, which remains relatively stable over 24 months. Given that the mental health of youth with gender dysphoria who are older is often poor, it is likely that puberty blockers prevent the deterioration of mental health.

*What’s known on this subject:* Puberty blockers are effective in halting endogenous puberty and prior research suggests improved mental health in youth with gender dysphoria. Few studies originate from the United States and most include older youth in later stages of puberty at initiation of blockers.

*What this study adds:* This is the largest longitudinal cohort of youth with gender dysphoria initiating medical intervention beginning with puberty blockers in early puberty to be followed in the United States. Youth demonstrated both stability and improvement in emotional and mental health over 24 months.

*Contributors Statement Page:* Dr. Olson-Kennedy conceptualized and designed the study, drafted the initial manuscript, and critically reviewed and revised the manuscript. Dr. Durazo-Arvizu carried out follow-up analyses and critically reviewed the manuscript. Drs. Garofalo, Rosenthal, Chan, Chen, Ehrensaft, and Hidalgo conceptualized and designed the study, and critically reviewed and revised the manuscript. Drs. Wang and Wong carried out the initial analyses and critically reviewed and revised the manuscript. Dr. Radix critically reviewed and revised the manuscript. All authors approved the final manuscript as submitted and agree to be accountable for all aspects of the work.

## Introduction

Transgender and non-binary (TNB) people identify as a gender that is misaligned with their phenotype. Many TNB individuals experience gender dysphoria—distress arising from this misalignment—and seek medical interventions to ease this distress. Often gender dysphoria arises or worsens with the onset of puberty.

Gonadotropin releasing hormone agonists (GnRHas) have been used in pediatric and adult populations since 1979 for the purpose of temporarily halting the production of gonadal sex steroids.[1] GnRHas in the United States are delivered most commonly via periodic injections (e.g., leuprolide acetate between 30 days and 6 months) or via an implant (histrelin acetate) inserted in the upper arm.[2] The implant is effective for 15-65 months.[3] After discontinuation of GnRHas the hypothalamic-pituitary-gonad axis will resume function after 6-12 months. [4]

Early studies from the Netherlands examining the mental and emotional health of TNB youth demonstrated that puberty blocker use halted the progression of endogenous puberty,[5] reduced symptoms of depression and anxiety and improved global functioning.[6] Following this model, The Endocrine Society and the World Professional Association of Transgender Health (WPATH) first recommended in 2009 and 2012, respectively, using GnRHas in early puberty to prevent the development of misaligned secondary sex characteristics associated with the progression of endogenous puberty.[7, 8]

Since these initial studies, additional evidence has demonstrated that GnRHas are associated with decreased suicidal ideation, behavioral and emotional problems, depressive symptoms and with better general functioning. [9–11] For example, in 2015, Costa et al. published findings from 200 youth with gender dysphoria demonstrating better global psychosocial functioning among youth starting pubertal suppression and psychotherapy concurrently compared with youth receiving psychotherapy alone.[9] In 2020, Van der Miesen et al. demonstrated that transgender adolescents administered puberty blockers had fewer emotional and behavioral problems than transgender youth just referred for assessment.[10] A recent study by McGregor et al. (2024) demonstrated that compared to TNB peers who had experienced a misaligned endogenous puberty, 40 youth who received puberty blockers in early stages of puberty (Tanner 2 or 3) reported less anxiety, depression, stress, and suicidal ideation and fewer total problems and internalizing difficulties.[11]

Based on the professional society recommendations noted above, puberty blockers are often used either as a temporizing measure to bring one’s gender identity into focus or as the initial stage in phenotypic gender transition, particularly when TNB youth access medical services in early adolescence. Previous research demonstrates that most but not all youth who initiate care with puberty blockers go on to use appropriate hormones for induction of aligned secondary sex characteristics.[6, 12, 13] Because the stages of care are individualized for each youth, time intervals between stages are quite variable, posing difficulties in studying puberty blockers as monotherapy.

In this study, we examine the mental and emotional health of a longitudinal, large and diverse cohort of youth initiating medical intervention predominantly in early puberty with GnRHas over 24 months.

## Methods

### Study Design and Participant Recruitment

Participants were enrolled between July 2016 and June 2019 as part of a larger study, the observational Trans Youth Care United States Study (TYCUS). Specific study procedures are detailed elsewhere.[14] Participants reported on outcomes of interest at baseline and subsequently every six months for two years post-initiation of medical treatment. This analysis examines a cohort of TNB youth initiating medical intervention with GnRHas. For each youth participant initiating GnRHas, one parent also completed questionnaires related to outcomes of interest. Ninety-five participants had baseline data, but one participant did not initiate GnRHas and was therefore excluded from analysis. Participants were eligible if they had a diagnosis of Gender Dysphoria; had onset of puberty to Tanner stage 2 or more, were already establishing medical care to undergo puberty suppression, and had the ability to read and understand English. Parent/Caretaker inclusion criteria included the ability to read and understand English. Youth participants were considered ineligible if they had prior use of GnRHas; had a diagnosis of precocious puberty (i.e., assigned male at birth and younger than 9 years or assigned female at birth and younger than 8 years); or pre-existing osteoporosis. All study procedures received approval from the Institutional Review Boards at participating institutions.

### Measures

#### Demographics and clinical characteristics

Participants self-reported demographic information, including age, race/ethnicity, gender identity, and designated sex at birth (DSAB). Racial/ethnic group differences were examined with Non-Hispanic White as the reference group. Recognizing the limitations in fully capturing the diversity of gender identities among TNB individuals, the sample was categorized and stratified based on DSAB to facilitate analysis. From the survey, we also assessed whether youth started GAHT by each time point during the 24-month study period. Finally, pubertal development based on breast development or testicular size was extracted from medical records.

#### Longitudinal Outcomes - Youth Report

### Beck Depression Inventory (BDI-Y)

Participants completed the BDI-Y, a 20-item self-report screener of depression symptoms over the previous two weeks. BDI-Y has good internal consistency, convergent validity, and intra-scale correlations; reliability coefficients range from *α* = .90 to *α* = .95.[15] Calculated T-scores reflect average (≤ 54), mildly elevated (55-59), moderately elevated (60-69), and extremely elevated (≥70) depressive symptoms.

### The NIH-Toolbox Emotion Battery

The NIH Toolbox Emotion Battery (NIHTB-EB) was employed to measure emotional health in a standardized manner. NIHTB-EB is comprehensive, assessing a wide range of both positive and negative aspects of social and emotional functioning. The scales included in the study were Self-Efficacy, Friendship, Loneliness, Emotional Support, Perceived Hostility and Perceived Rejection. The T-scores reported here were uncorrected. According to the NIH, uncorrected scores provide a glimpse of the given participant’s overall performance when compared with the population of the US as determined by the Census. The uncorrected score may be most useful when trying to gauge one’s overall level of functioning, not in the context of age, gender, or other demographic factors. (NIH Toolbox Scoring and Interpretation Guide for the iPad © 2006-2016 National Institutes of Health and Northwestern University). Our previous research supports NIHTB-EB as a suitable assessment of TNB individuals.[16]

### Suicidality

Suicidality (both lifetime and past 6 months) was assessed with up to six “yes” or “no” items adopted from the Middle School Youth Risk Behavior Survey, the largest national survey of youth behavior and mental health.[17] Participants who endorsed lifetime suicidality were subsequently asked about suicidality in the past 6 months.

#### Longitudinal Outcomes - Parent Report

### Child Behavior Checklist (CBCL)

CBCL was used to capture parent/guardian reports of youth mental health functioning at baseline, 12 months and 24 months. The CBCL is a well-validated, psychometrically sound measure of behavioral and emotional problems in youth ages 6 to 18 years. The scale measures eight syndrome scales (anxious depressed, withdrawn depressed, somatic complaints, social problems, thought problems, attention problems, rule breaking behavior and aggressive behavior) and two higher-order factors (internalizing and externalizing problems).[18] The results are reported as T-scores, with scores >63 indicating that additional investigation may be warranted in clinical settings. CBCL scores were calculated using gender neutral norms as outlined by ASEBA. (Achenbach, T.M., & Ivanova, M.Y. (2022). Guide to Gender-Inclusive Assessment Using the ASEBA. Burlington, VT: University of Vermont, Research Center for Children, Youth, & Families.)

### Statistical Analysis

Latent Growth-Curve Models (LGCM) within the framework of Structural Equation Modeling (SEM) were employed to analyze the data using Mplus version 8.5.[19] The LGCM estimated means and variances of parameters at baseline (i.e., intercept factor) and changes over time (i.e., slope factor). Bayesian estimation with Markov chain Monte Carlo resampling was employed, suitable for the moderate sample size of the study.[20] Model convergence[19] was assessed using the Gelman-Rubin potential scale reduction factor, with values approaching 1 indicating convergence. Trace plots were examined to ensure model fit, confirming successful model convergence with PSR values ranging from 1.01 to 1.03. The unconditional LGCM characterized changes without covariates, while the conditional model incorporated time-independent variables including age, DSAB, Tanner stage at baseline, and racial/ethnic group membership. We also included a variable reflecting whether participants had started GAHT at each time point during the study period as a time-varying covariate. We employed the following model selection process: (1) Unconditional model; (2) Added a GAHT start indicator as a time-dependent covariate; (3) A conditional model that included time-independent covariates. This model was specified so that age at baseline and DSAB were assumed to be related to the slope. This modeling approach resulted in final models containing all time-independent covariates.[21] The patterns of missing data were examined, employing Full Information Maximum Likelihood methods for the estimation of model parameters when data is missing at random.[22]

## Results

Ninety-four participants initiating GnRHas ranged in age from 8-16 years M (SD) = 11.20 (1.46) and included a predominance of Non-Hispanic White participants (n = 53; 56.4%). See Table 1 for additional demographics. Slightly more than half of participants were designated male at birth (n = 49). Eighty participants were in early puberty (Tanner stage 2 or 3) at baseline. Of the analytic sample, eleven participants started gender-affirming hormone treatment (GAHT), estradiol or testosterone within the first 12 months after GnRHa initiation, and an additional 20 participants started GAHT between 12- and 24-months. For those starting GAHT, average time to GAHT start was 1.18 years (range 0.17-1.92 y).

**Table 1.**
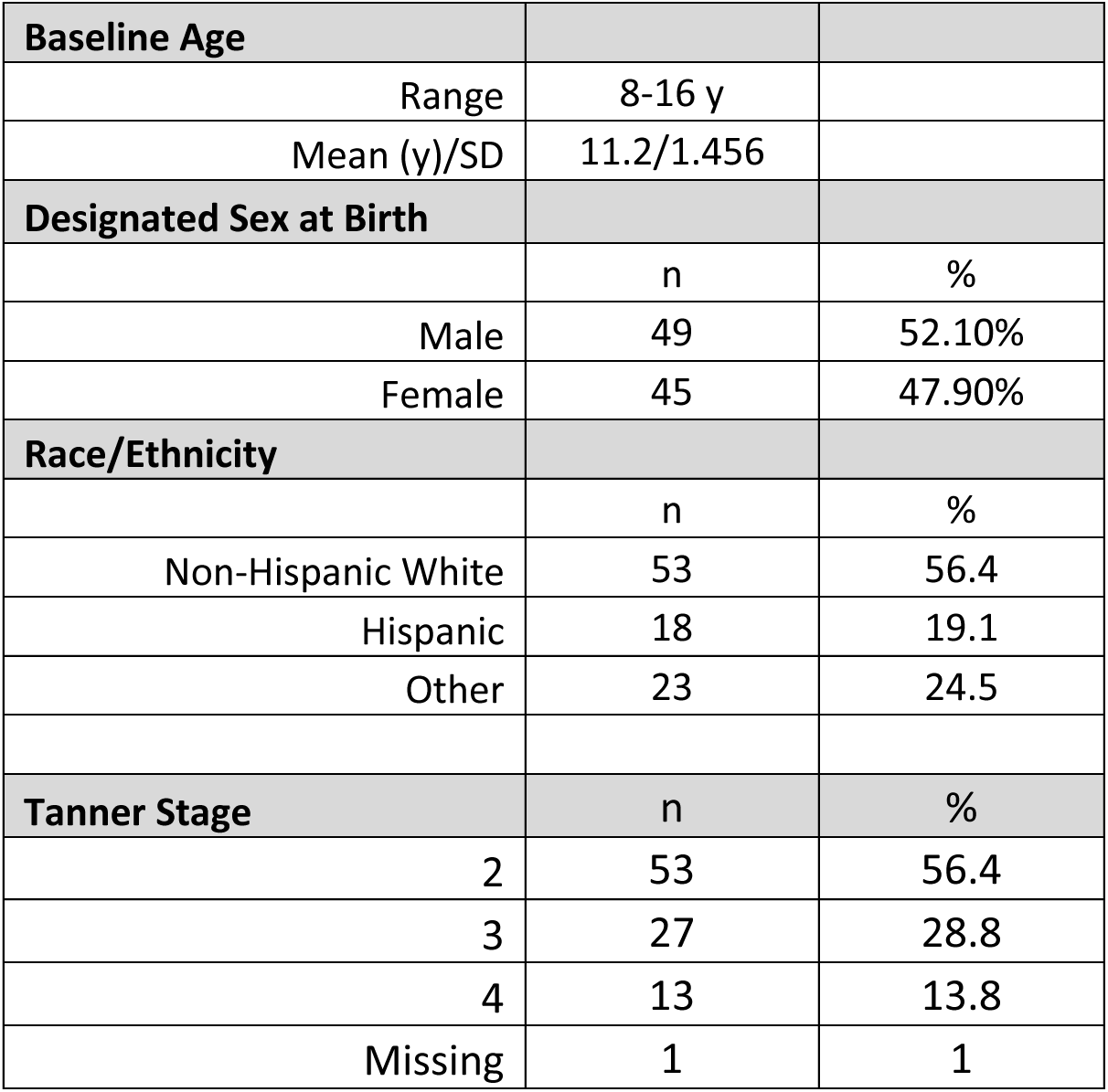
Demographics.

| **Baseline Age** |  |  |
| --- | --- | --- |
| Range | 8-16 y |  |
| Mean (y)/SD | 11.2/1.456 |  |
| **Designated Sex at Birth** |  |  |
|  | n | % |
| Male | 49 | 52.10% |
| Female | 45 | 47.90% |
| **Race/Ethnicity** |  |  |
|  | n | % |
| Non-Hispanic White | 53 | 56.4 |
| Hispanic | 18 | 19.1 |
| Other | 23 | 24.5 |
| **Tanner Stage** |  |  |
|  | n | % |
| 2 | 53 | 56.4 |
| 3 | 27 | 28.8 |
| 4 | 13 | 13.8 |
| Missing | 1 | 1 |

### Youth-Reported Outcomes

#### Depression BDI-Y

Self-reported depression symptoms did not change significantly over the 2-year follow-up period in both the conditional and unconditional growth curve models. (Table 2a) At baseline, 72% percent (n= 63) of participants had average BDI-Y scores, 10% (n=9) mildly elevated, 10% (n=9) moderately elevated, and 8% (n=7) severely elevated. At 24-months (n=59), 75% (n=42) of participants had average BDI-Y scores, 7% (n=4) mildly elevated, 14% (n=8) moderately elevated, and 9% (n=5) severely elevated scores (Table 5). Means across time points are displayed in Table 2a. In the conditional model (Table 3), age at baseline had an impact on baseline BDI-Y scores, but no covariates were associated with change over time. A one-year increase in baseline age is associated with an increase of 3.78 (95% CI: 1.99-5.57) on the baseline BDI-Y T score.

**Table 2.**
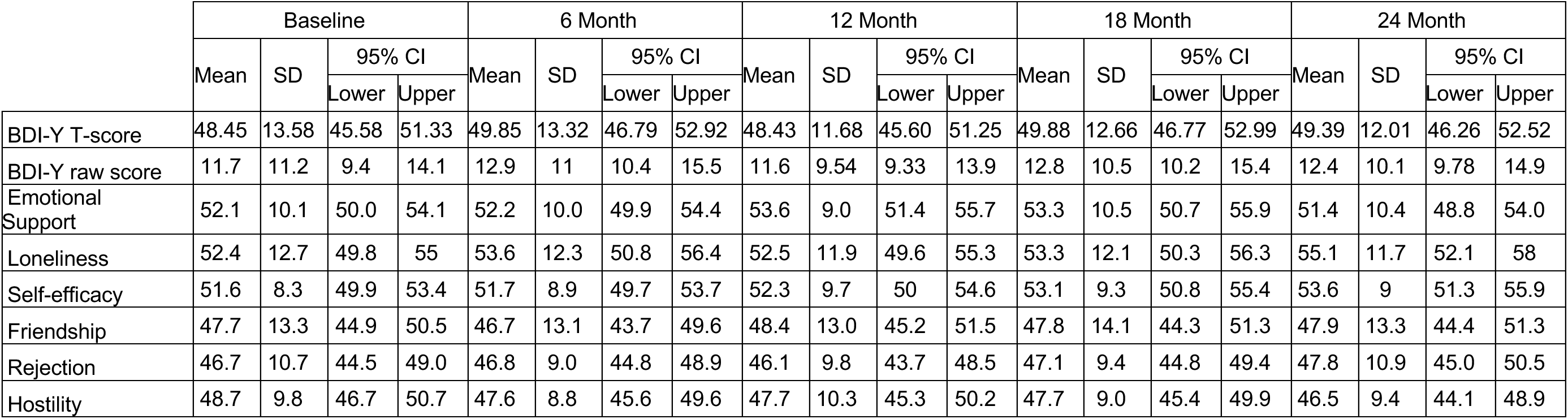
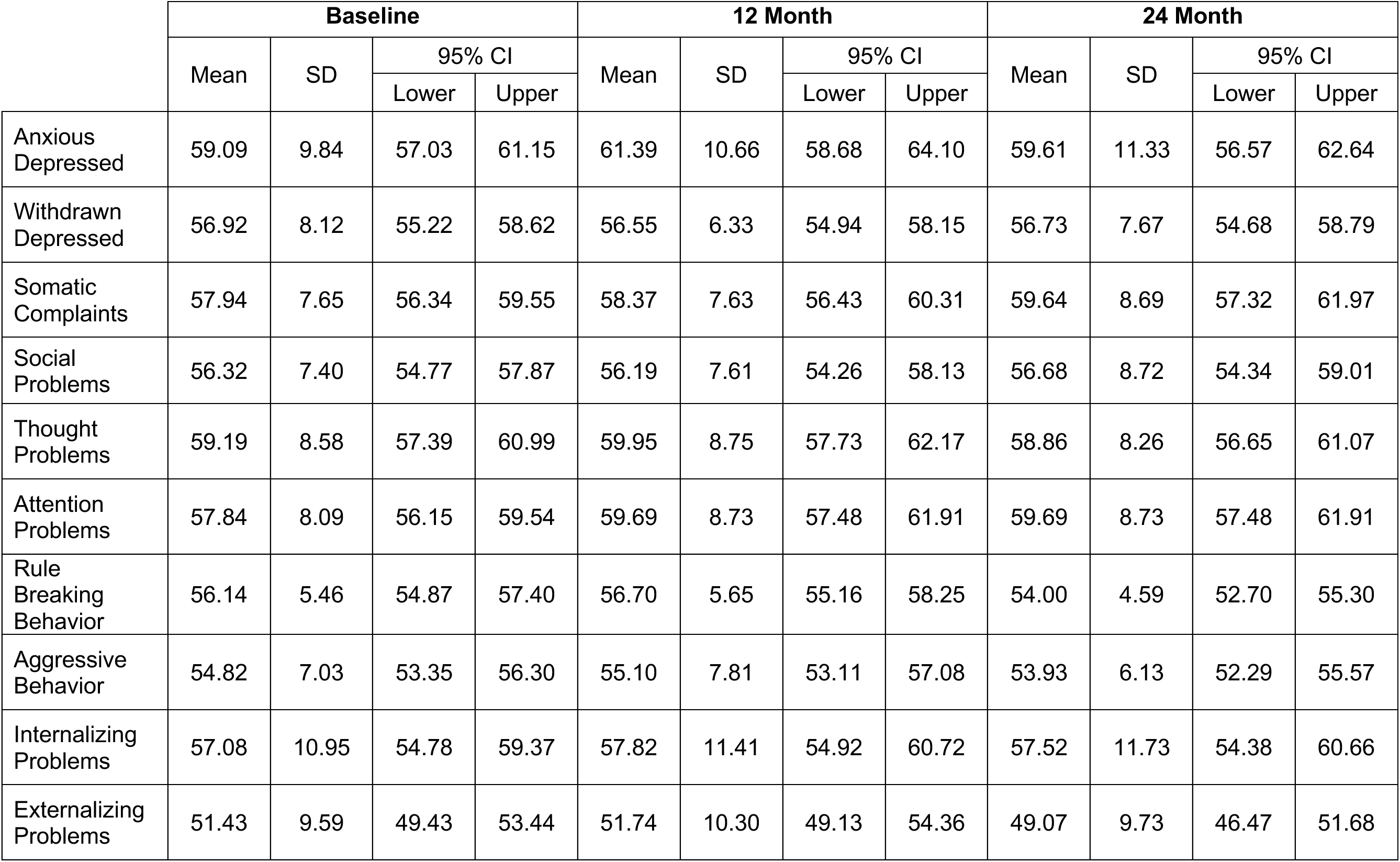
a. Descriptives of BDI-Y and NIHTB-EB b. Descriptives of CBCL Outcomes

**Table 4:**
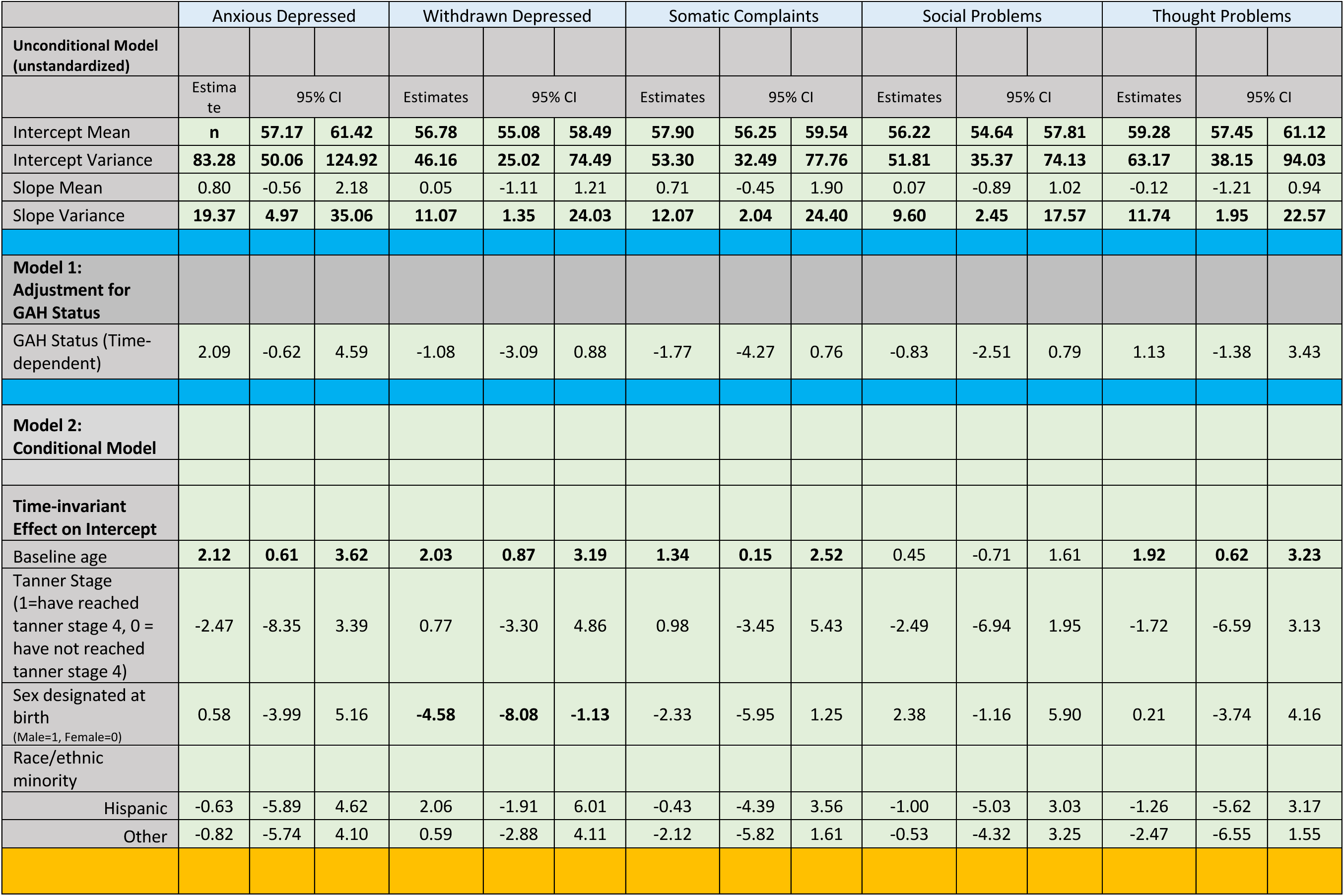
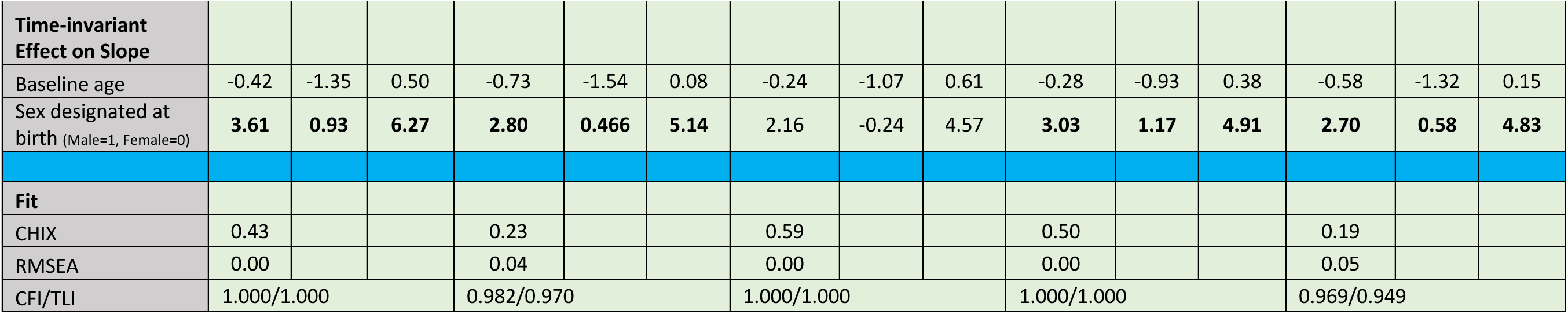
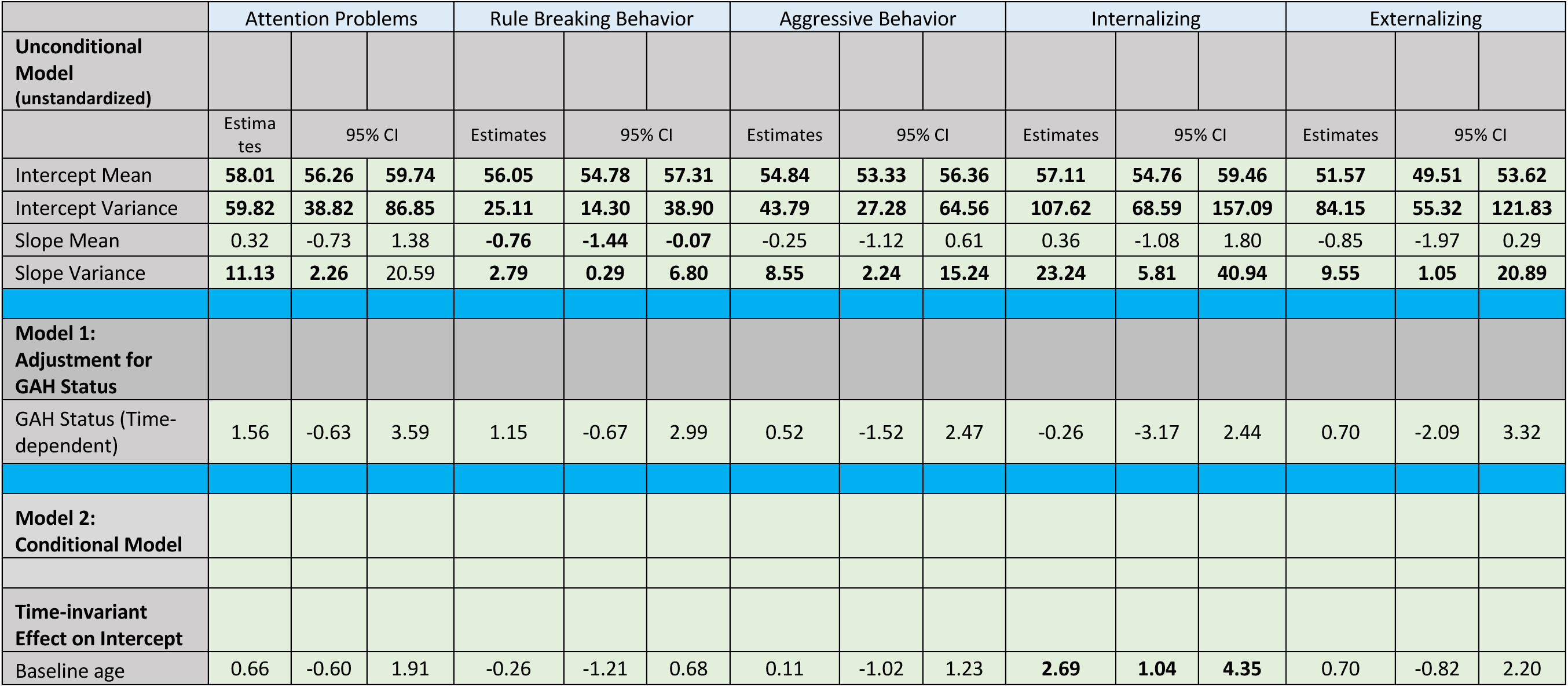
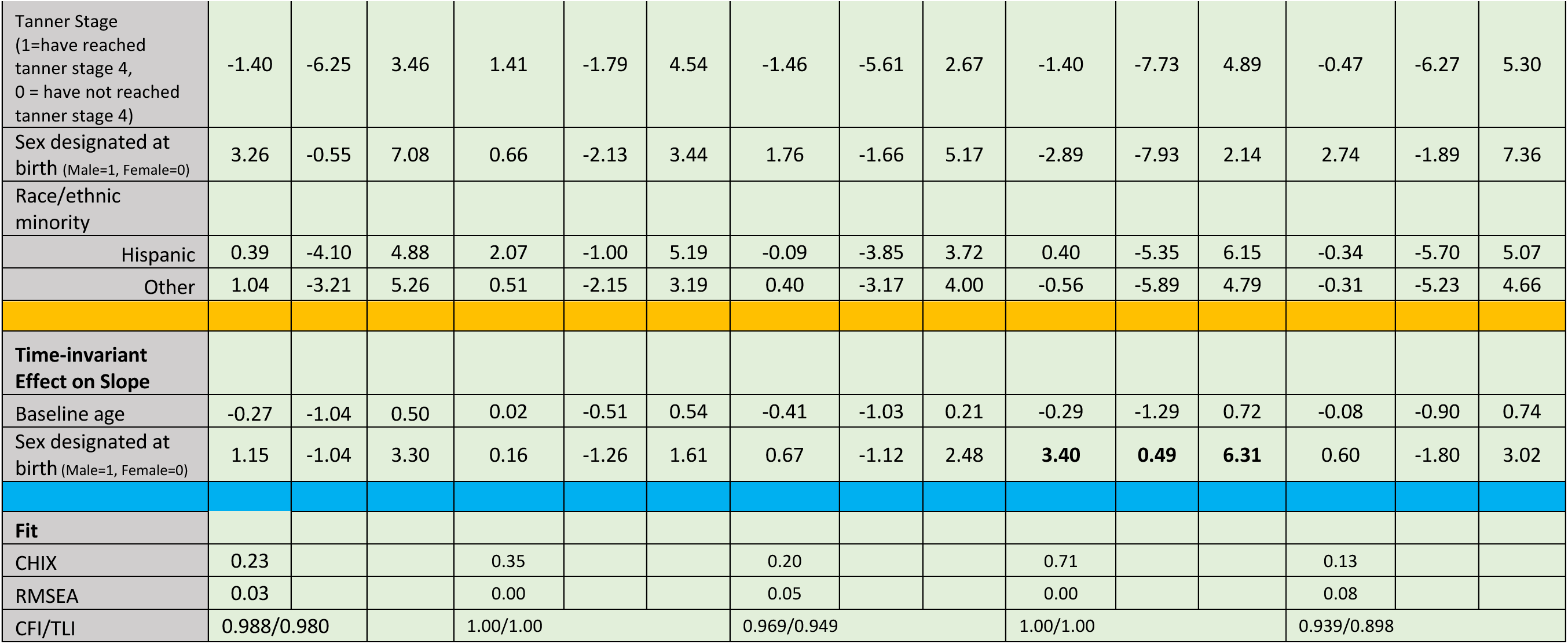
LGCM CBCL.

**Table 5:**
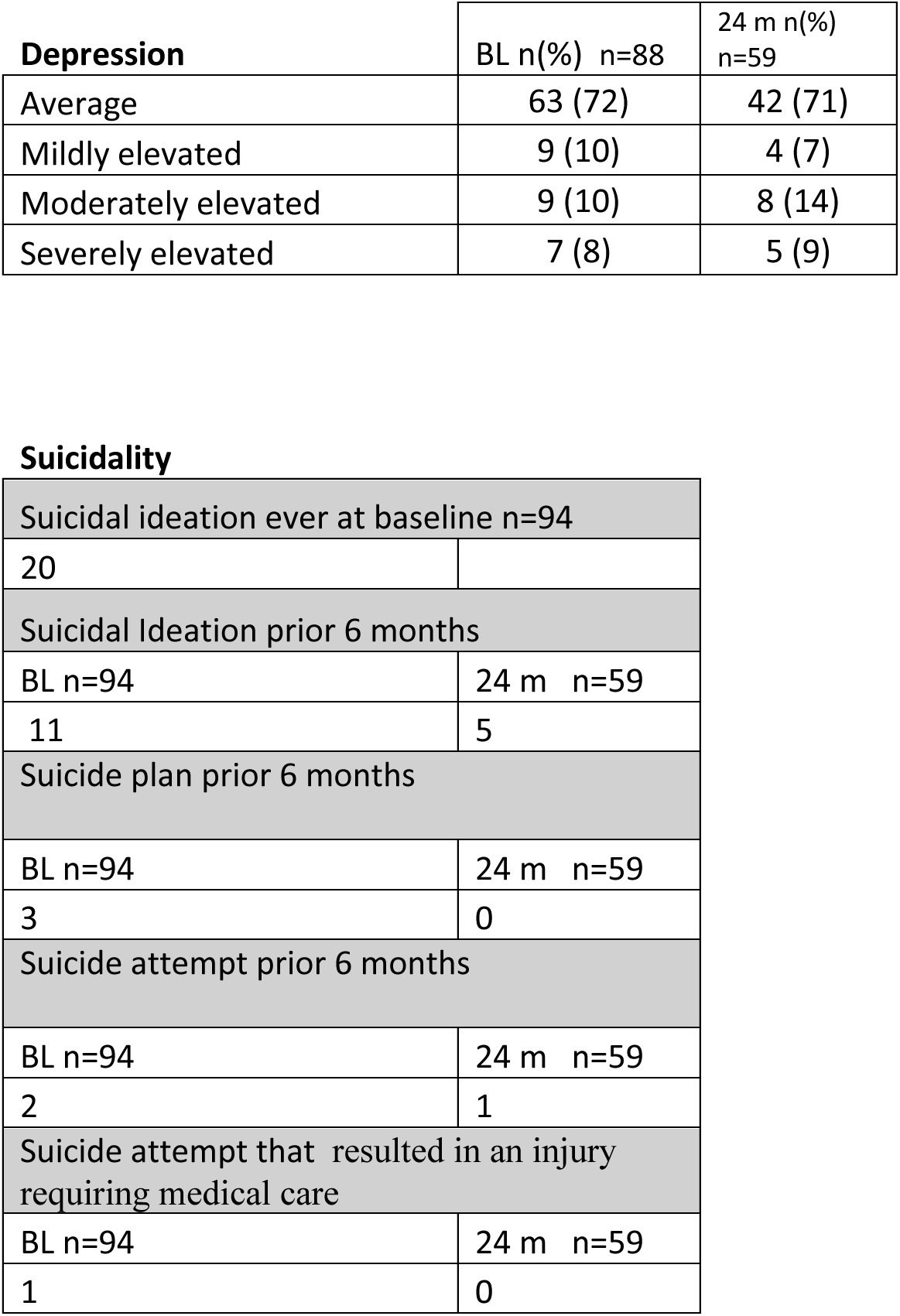
Depression and Suicidality.

| **Depression** | BL n(%) n=88 | 24 m n(%) n=59 |
| --- | --- | --- |
| Average | 63 (72) | 42 (71) |
| Mildly elevated | 9 (10) | 4 (7) |
| Moderately elevated | 9 (10) | 8 (14) |
| Severely elevated | 7 (8) | 5 (9) |

Suicidality
|  |  |
| --- | --- |
| Suicidal ideation ever at baseline n=94 |  |
| 20 |  |
| Suicidal Ideation prior 6 months |  |
| BL n=94 | 24 m n=59 |
| 11 | 5 |
| Suicide plan prior 6 months |  |
| BL n=94 | 24 m n=59 |
| 3 | 0 |
| Suicide attempt prior 6 months |  |
| BL n=94 | 24 m n=59 |
| 2 | 1 |
| Suicide attempt that resulted in an injury requiring medical care |  |
| BL n=94 | 24 m n=59 |
| 1 | 0 |

##### Suicidality

At baseline, 20 participants reported ever experiencing suicidal ideation, 11 participants endorsed suicidal ideation in the prior 6 months, 3 participants had made a suicide plan in the past 6 months, and 2 participants reported a suicide attempt in the past 6 months, one of which resulted in an injury requiring medical care. At 24-month follow-up, 5 participants endorsed suicidal ideation in the prior 6 months, no participants had made a suicide plan in the past 6 months, and 1 participant reported a suicide attempt in the past 6 months which did not result in an injury requiring medical care. There were no suicide deaths over the 24-month time period. (Table 5)

#### NIHTB-EB

The unconditional growth model for NIHTB-Emotional Battery demonstrates no significant change in these self-reported symptoms across the study period (Table 3). At no time points were the mean scores in a clinically concerning range (>63) for any of the Emotional Health NIH toolbox domains (Table 2a).

The conditional model incorporated covariates to assess their impact on both the initial scores and the trajectories of the battery of emotion measures.

**Baseline Age** significantly impacted loneliness and rejection scores at baseline. A one-year increase in baseline age is associated with an increase of 3.03 (95% CI: 1.29-4.76) on the loneliness score, and an increase of 1.55 (95% CI: .08-3.02) on the perceived rejection score. Baseline age had no impact on slope on any of the NIHTB-EB constructs.
**Tanner stage, Designated Sex at Birth, Race/Ethnic Minority Status** had no impact on baseline values or rates of change for any of the NIHTB-EB measures.
GAHT Status at a visit had no bearing on any of the NIHTB-EB T-scores at the same visit.

### Parent Reported Outcomes

#### Longitudinal Changes in CBCL

The unconditional LGCM (Table 4) demonstrated no significant changes in reported anxious/depressed, withdrawn/depressed, somatic complaints, social problems, thought problems, attention problems, aggressive behavior, internalizing problems or externalizing problems in the overall sample over the 24-months after initiating GnRHa. Participants demonstrated a significant decrease in rule-breaking behaviors, β (mean rate of decrease over 24 months) −0.76, 95% CI: (−1.44, −0.07). Means across time points are displayed in Table 2b.

### Effects of Covariates on CBCL outcomes

The Conditional Model (Table 4) assessed effects of covariates in predicting initial values and slopes of outcomes of interests.

**Age at baseline** was a significant predictor of higher baseline scores, with older youth exhibiting higher reported anxious/depressed β= 2.12 (95% CI: 0.61-3.62) point/year, withdrawn/depressed β=2.03, 95% CI: (0.87, 3.19)] points/year, somatic complaints β=1.34, 95% CI (0.15-2.52) points/year, thought problems β= 1.92, 95% CI: (0.62, 3.23) points/year, and internalizing problems scores β= 2.69, 95% CI: (1.04, 4.35) points/year. Age at baseline had no impact on change in these constructs over time.
**Tanner stage** at initiation of GnRHa did not predict initial values or slopes in CBCL scores.
**Designated sex at birth** was a significant predictor of baseline withdrawn depressed scores β= −4.58, 95% CI: (−8.08, −1.13), with those assigned male at birth having significantly lower baseline mean withdrawn depressed scores than those assigned female at birth. In addition, designated sex at birth was a significant predictor of change in several domains of the CBCL. Specifically, those assigned male at birth compared to those assigned female at birth exhibited faster change in scores for anxious/depressed at a rate difference of β= 3.61, 95% CI: (0.93,6.27) points/year, withdrawn/depressed at a rate difference of β= 2.8 CI: (0.466, 5.14) points/year, social problems at a rate difference of β= 3.03, 95% CI: (1.17,4.91) points/year, thought problems at a rate difference of β= 2.70 CI: (.58,-4.83) points/year and internalizing symptoms at a rate difference of β= 3.4, 95% CI: (0.49, 6.31) points/year.
**Minoritized race/ethnicity** did not predict initial values nor slope for any of the reported CBCL domains.
**Intercept and Slope Variances:** The estimated values and 95% confidence intervals suggest a high degree of diversity in the baseline scores (intercepts) and rates of change (slopes).

## Discussion

Prior findings suggest that youth with gender dysphoria who have access to puberty blockers either demonstrate improved mental health or experience neutral effects, potentially preventing worsening mental health related in part to gender dysphoria.[9–11, 23] The results presented here are consistent with the prior literature and demonstrate that the mental health of youth as reported by both themselves and their parents/guardians is relatively stable from baseline over 24 months after starting medical intervention with GnRHa.

Depression commonly accompanies gender dysphoria and has been routinely reported as higher among youth with gender dysphoria than the population of youth at large.[24, 25] In the Youth Risk Behavioral Health Survey (YRBS) from 2021, 23.5% of middle school students reported that their mental health was not good “most of the time” or “always.” [26] While it is not certain how “most of the time” or “always” translates to BDI-Y moderate or severe ranges, it is reasonable to assume these offer acceptable proxies, suggesting that at both baseline (18%) and at 24 months (22%), the proportion of our sample with poor mental health is no greater than that described in the general population.

National studies also report that among middle school youth, suicidal ideation, plans and attempts have continued to increase over time. In 2019, 22.3% of middle school youth reported ever thinking about suicide, 14.7% reported ever making a suicide plan, and 9.3% reported ever attempting suicide in their lifetime.[27] Suicidality is a major concern for all youth, but particularly among sexual and gender minoritized youth including those who identify as TNB.[28] Among our participants at baseline lifetime suicidal ideation, plan and attempts were comparable to those of the general population, and after 24-months following initiation of medical intervention with GnRHa, were lower than the national average.

Among the constructs assessed within CBCL, most showed no significant change over time. It is important to note that the means reported across the entire sample were not in a clinically concerning range at any time point. Rule-breaking behavior demonstrated a statistically significant decline (improvement) over time. Because the mean reported for rule-breaking behavior was not clinically concerning at any time point, the clinical significance of this change is unclear.

### Strengths and Limitations

A primary strength of the study is the prospective, longitudinal design with multiple assessment points across time. This study used widely validated instruments to examine emotional and psychological health reported by both youth and their parents/guardians as respondents. Finally, this study included a more racially and ethnically diverse sample of participants, the majority of whom were in early puberty (Tanner stage 2/3), in comparison to other research examining the impact of puberty suppression. Study limitations include that more than half of the participants had initiated gender-affirming hormones over the 24-month follow up period, limiting our ability to examine puberty suppression as monotherapy. Another limitation is that all four sites are located in urban areas, so findings are not necessarily generalizable to youth in more rural areas of the country. Finally, all participants were able to access care and participate in the study, which indicates some level of parental support. Parental support has been found to contribute positively to mental health among all adolescents;[29, 30] therefore, these findings may not be generalizable to youth who have minimal or no parental support.

## Conclusion

The mental health and well-being of youth with gender dysphoria after initiating medical intervention is of clinical interest to TNB youth, their providers, and their parents/guardians. Among this larger sample of youth initiating medical intervention first with GnRHas to address gender dysphoria, the average mental health as reported by both the youth and parents remains in a clinically non-significant range over two years. The findings presented here add to the existing literature on mental health among youth with gender dysphoria undergoing gender-affirming medical treatment.

## Data Availability

All data produced in the present study are available upon reasonable request to the authors

## Abbreviations

BDI-Y: Beck Depression Inventory – Youth
CBCL: Child Behavior Checklist
DSAB: designated sex at birth
GAHT: gender affirming hormone therapy
GnRHa: gonadotropin releasing hormone agonist
NIHTB-EB: The NIH Toolbox Emotion Battery
TYCUS: Trans Youth Care United States Study
TNB: Transgender and non-binary
WPATH: World Professional Association of Transgender Health

